# Recovery in women with severe mental illness in long-stay facilities: Protocol for an ethnographic & art-based inquiry

**DOI:** 10.1101/2025.04.03.25325179

**Authors:** Kantadorshi Parashar, Poornima Bhola, Krishna Prasad Muliyala, Sarah Pinto

## Abstract

**Background:** Recovery and rehabilitation in severe mental illness (SMI) often involve periods of institutionalization, which can sometimes extend longer than anticipated. Understanding the lived experiences and recovery narratives from context-specific, insider perspectives in such long-term settings is crucial for shaping care and support that aligns with individual aspirations. This article outlines the protocol of a research study designed to conduct an ethnographic and art-based inquiry into recovery in women with SMI residing in long-stay facilities in India.

**Method:** The study will have a multi-site ethnographic and art-based inquiry design to be conducted in 3 long-stay facilities (comprising of psychiatric hospitals, NGO homes, and government-run state homes) across different geographical locations. The researcher will spend between 20 to 25 days for participant observation in each setting, writing field notes, and conducting ethnographic interactions with staff and service-users. An organization information sheet will be used to record the various facilities available in the setting. A sub-sample of 24 service-users will be approached for an in-depth interview about their lived experience of recovery, and assessments using the Clinical Global Impressions scale (CGI), pictorial recovery tool Swasthya Labh Saadhan (SLS), and Cantril’s ladder. Art-based inquiries using a list of prompts will be carried out over 10 sessions in an open group of interested service-users. The data will be analysed using both qualitative and quantitative methods.

**Discussion:** Findings from the proposed study will build an understanding of the multiple narratives of recovery that are rooted in the unique personal and cultural contexts of service-users. Additionally, understanding various kinds of long-stay facilities, housing environments, and organizational practices will help us map care practices and infrastructures that promote recovery-oriented practices.

## Introduction

The narrative of recovery in severe mental illness is increasingly viewed as a process rather than as an outcome, with lived experience at the core. Each person’s journey through and with mental illness is unique and should be viewed beyond the singular goal of symptom elimination. These narratives bring structure to events and impose order on chaos, thereby creating meaning.^1^ The meanings of recovery enable individuals to be seen beyond the labels of their diagnoses or the challenges they entail, shifting the focus to their strengths, potential, passions, and aspirations, even within the constraints posed by the illness.^2,3^ The process of recovery unfolds across different physical spaces-homes, hospitals, and institutional care facilities. Some individuals with severe mental illness (SMI) experience prolonged stays in institutional settings, with a global median of 18% of people in psychiatric facilities remaining there for a year or more.^4^ The reasons for such prolonged stay could be the absence of family members, lack of financial support for independent living, helplessness, abandonment by family, or done by choice/voluntarily.^5,6^ Life within these long-stay institutions is a chronicle of both vulnerability and resilience. Despite being one of the most profound manifestations of social exclusions due to mental illness,^6^ accounts of shared living, surrogate families, cohesiveness, and recovery journeys also persist in institutions.

In India, women make up 54.3% of the population staying in state-run psychiatric facilities for a year or more, with the majority experiencing severe mental illness (SMI).^6^ Positioned at the intersections of multiple social identities, power relations, and experiences, their realities and perspectives must be taken into account to determine the nature of support they receive. To understand the relationship between the idea of recovery and what it means in actual practice, we need to ask questions to different people in the local context. Such questions, when rooted in research values of justice, dignity, and equity, allow us to tap marginalized and silenced voices. Studies^5,7^ that have delved into the living conditions, needs, and possible paths of intervention for women with SMI in long-stay facilities have called attention to revising care practices at multiple levels. However, there is also a need for immersive, participatory approaches that incorporate insider perspectives, practice-near accounts, and delineate information rooted in the context and sub-culture. Long-stay experiences vary across types of facilities-such as, a government-run State Home would have women both with and without mental illness staying together, while a tertiary mental health institution will have provisions exclusively for women with mental illness. The routine and restrictions also vary along with the spectrum of symptom severity in these facilities. For instance, tertiary settings have acute and chronic cases staying together, with routine visits from nurses and other mental health professionals, with most janitorial and housekeeping services outsourced. A home run by the government has occasional such professional visits, with most cases being chronic in nature and diverse interpersonal dynamics as service-users contribute to several chores within the facility. Both settings, however, have women who have attained symptomatic recovery but are restricted to the physical boundaries due to different reasons, while another proportion remains with treatment-resistant symptomology and limited socio-occupational functioning. The lived experiences of recovery and meaning-making may only be adequately understood with research methods closer to the context and its experience. The physical structures and care-practices that aid recovery from mental illness in long-stay facilities require to be re-imagined in a way that there is a change from a control and management perspective to one that is choice-based across multiple domains.

A researcher’s gradual entry into the participants’ physical space of experience, routine, conversations, and everyday processes makes it possible to tap into the discourse of recovery and calls for incorporating these voices through modes of expression apart from language. Arts-based and participatory research methods can help capture these complex narratives and enable marginalized voices to be heard beyond barriers of language, literacy, symptom severity, and speech deficits. Such methods also support researchers in building rapport and trust with participants, allowing sensitive topics to emerge in a non-threatening way.^9^ This aligns with a recovery-oriented mental health model that prioritizes collaboration, inclusivity, and self-directed recovery, giving women with SMI the opportunity to be seen as experts in their own lives and experiences. A recovery-oriented approach necessitates a shift from services based on paternalistic and either protective or restrictive approaches to ones that value the aspirations and needs of service users and regard them as experts on decisions related to their well-being. Such practices emphasize hope, social inclusion, community participation, personal goal setting, and self-management, and they promote a partnership between people accessing mental health services and mental health professionals.^10^

The proposed study will employ a multi-centric, mixed-methods approach, combining ethnographic and arts-based methods to gather nuanced, insider perspectives on recovery among women in long-stay settings. This approach will help bridge gaps in understanding by revealing context-rich data crucial for shaping inclusive, recovery-oriented policies in India’s institutional settings.

### Objectives of the Study

The primary aim of the study is to conduct an ethnographic and art-based inquiry into recovery in women with SMI in long-stay facilities, with the following objectives:

1. To understand the meaning/experience of recovery among women with SMI in long-stay facilities.
2. To explore the housing environment and recovery-oriented practices for women with SMI in long-stay facilities.

## Materials and Methods

### Study Design and Setting

The study will have a multi-site ethnographic and art-based inquiry design involving participant observations, in-depth interviews, and art-based interactions. It will be conducted in three types of long-stay facilities: (i) psychiatric hospitals, (ii) non-governmental organization (NGO) homes, and (iii) government-run state homes for women; across different geographical locations of India. The facilities will be sampled based on convenience and pragmatic factors such as language, geographical location, and travel feasibility.

### Operational Definitions

#### Long-stay

Any period of stay in a long-stay facility of one year or more, as outlined in the National Strategy for Inclusive and Community Based Living for Persons with Mental Health Issues.^6^

#### Long-stay facilities

Any establishment where persons with long-term care needs can stay and access health and welfare benefits away from family, relatives, or friends. For the purpose of the current study, we will look at three kinds of facilities:

i. Psychiatric hospitals: Tertiary centers which specialize in the treatment of mental disorders, varying in size and grading,^11^ and have indoor facilities for patients without attendants.
ii. NGO homes: NGO refers to different categories of entities that operate not to obtain financial gain and also do not belong to the government sector. The current study will include registered NGOs that provide residential care services to women with mental illness.
iii. Government run State homes for women: Homes run by respective State governments for women who do not have any shelter, and includes women from diverse background, living under one roof.

#### Severe mental illness (SMI)

A mental, behavioural, or emotional disorder resulting in serious functional impairment, which substantially interferes with or limits one or more major life activities.^12^ For the purpose of this study, SMI will be inclusive of ICD-10^13^ diagnoses of F20-F29 (Schizophrenia, schizotypal and delusional disorders), F31 (Bipolar affective disorder), F32.3 (Severe depressive episode with psychotic symptoms), or F33 (Recurrent Depressive disorder), as diagnosed and documented by the treating team in the long stay facility.

#### Recovery

A deeply personal journey of varying narratives involving development of meanings beyond the catastrophic effects of mental illness.^2^ According to Swasthya Labh Saadhan (SLS) pictorial recovery tool,^14^ recovery involves i) taking care of oneself, ii) being addiction free, iii) being spiritually engaged, iv) having fun, v) being an active family member, vi) being a friend, vii) contribute to the household, and viii) being an active community member.

#### Recovery-oriented practices

An approach towards mental health care that promotes a partnership between patients and mental health professionals (MHPs), across dimensions of: i) design, ii) evaluation, iii) leadership, iv) management, v) integration, vi) comprehensiveness, vii) consumer involvement, viii) cultural relevance, ix) advocacy, x) training, xi) funding, and xii) access.^15^

#### Housing Environment

Housing environments refer to physical attributes of an environment that are tangible and observable, such as open space, number of rooms, occupants, and social and psychological attributes, such as experiences in forging relationships, feeling of belongingness, and safety;^16^ to be studied through organization information sheet and participant observation.

#### Ethnography

Ethnography refers to the qualitative research method in which a researcher studies a particular social/cultural group with the aim to better understand it, by actively participating in the group to gain an insider’s perspective of the group and create an account of the group based on this participation, interviews with group members,^17^ and an analysis of field notes of participant observations which will be written using the observation guide

#### Art-based inquiry

Art-based inquiry refers to a set of tools that draw on representational forms rooted in tenets of creative arts in order to address research questions.^18^ For the current study, representational forms of, but not limited to, paintings, drawings, clay work, and collages will be used. All of these will be a mode of inquiry and research tool, and not an intervention.

### Study Tools

#### 1. Organization information sheet

An information sheet to gather details about the long-stay facility will be developed. It will include details such as total number of service-users, number and nature of staff, number of rooms, key services that are provided, period since when the organization is providing services, collaborations with other organizations. This information will be collected through participant observation, and also from staff in the particular centre who are in leadership roles for at least 1-year duration. Informed written consent will be sought from them before seeking such information.

#### 2. Participant Observation

Ethnographic field research involves the study of groups and people as they go about their everyday lives.^19^ Hereby, participant observation will be carried out by the first author, who is female, in each of the long-stay facilities with service-users who provide informed written consent for the study.

The researcher will spend between 20 and 25 days in each long-stay facility. As per Jeffrey & Troman^20^, an ethnographic approach in a compressed mode can involve a short period of intense ethnographic research in sites from a few days to a month. In such an approach, ethnography captures the dynamics of a context, documenting the visible and less tangible social structures and relations. Observational field notes, a key element of the data, will be collected by spending significant time in the research environment, attentively capturing the routines, tensions, and disruptions within the research site.

An observation guide will be used as an anchor point for jotting field notes to ensure that different dimensions of a situation are adequately observed and noted. This guide will be developed by all researchers in consultation with each other, which will then be content validated by five experts (See Table 1 for selection criteria of content validation experts).

**Table 1.**
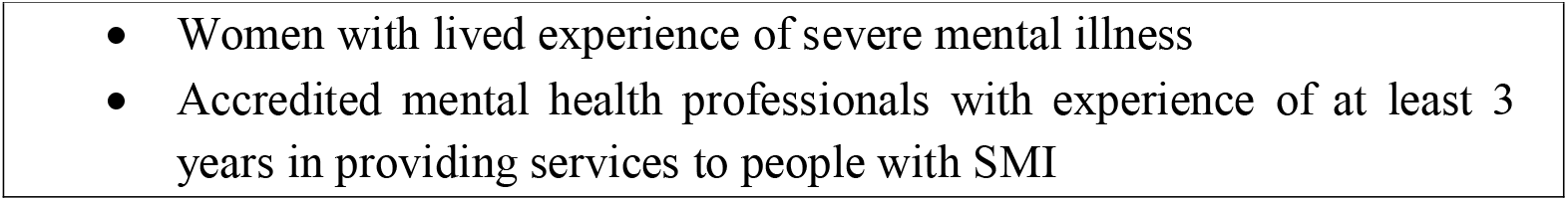
Selection Criteria for Content Validation Experts.

To ensure adequacy of data and address data triangulation, the following cautions will be maintained:

i. Organizing information *in situ*^20^: All observations and thoughts recorded in field notes will be organized at the end of every day and contain full details of all observations made in the day.
ii. To keep a framework of awareness to bias and inclusivity of different narratives, the research will also engage in writing personal reflections and positionality statement.

Apart from the direct observations that the researcher makes, in the process of acquainting with the people involved, the researcher will also interact with key persons in the centres which may involve mental health professionals, case managers, nurses, health care workers, personal assistants, administrative staff, superintendents of the institute, and NGO representatives. The inputs from these interactions will be incorporated in the field notes.

In the current study, field notes will involve jottings and initial impressions; description of things particularly of interest to the research questions, based on the observation guide; writing what people in the setting experience and react to as ‘significant’ or important, with additional impetus on how routine actions in the setting are organized and carried out; and reflection of what the researcher has thought, felt, and learned when making observations.^19,21^

#### 3. In-depth interviews and assessments

In-depth interviews and assessments will be carried out with the goal of obtaining a detailed and rich understanding of the participant’s lived experience and conceptualization of recovery. The participants will be selected as a purposive sub-sample (n=24) from the cohort of women who are being observed in the long-stay facility. The interviews and assessments will thus complement participant observation and will be conducted by the researcher in private rooms in field sites in between participant observations. The selection criteria of participants will be:

##### Inclusion criteria for in-depth interviews and assessments

i. Female service users with a diagnosis of F20-F29 (Schizophrenia, schizotypal and delusional disorders), F31 (Bipolar affective disorder), F32.3 (Severe depressive episode with psychotic symptoms), or F33 (Recurrent Depressive disorder) as per ICD-10^13^
ii. Age 18 to 60 years
iii. Staying in the long-stay facility for at least 1-year duration
iv. Working knowledge of Hindi or English or Assamese or Bengali
v. Has the capacity to consent for participation in the study.

##### Exclusion criteria for in-depth interviews and assessments

i. Co-morbid Intellectual or Developmental Disability on clinical impression/records
ii. Current substance dependence, except nicotine
iii. History of Traumatic Brain Injury TBI or organic condition that would interfere with participation in the study

The in-depth interviews and assessments will involve:

i. Socio-demographic and clinical data sheet: A semi-structured socio-demographic and clinical data sheet will be prepared by the researcher to record the details of the service-users who provide informed written consent for in-depth interview and assessments. It will include information such as age, gender, education, occupation, marital status, and clinical history, including, but not limited to, psychiatric diagnosis, history of admission into the long-stay facility, intervention goals, information about efforts for community reintegration, and reasons for a prolonged stay. This will also include a check-list of legal identity documents available to women in the long-stay, indicating proof of identity, address, and disability, such as an Aadhar Card and Election Commission ID card. The tool will be administered by the researcher.
ii. Clinical Global Impressions Scale (CGI)^22^: The CGI provides an overall clinician-determined summary measure that takes into account all available clinical and psychosocial information.^23^ The CGI is a 3-item observer-rated scale that measures illness severity (CGIS), global improvement or change (CGIC), and therapeutic response. The tool will be administered by the researcher only to rate the illness severity domain (CGIS); other domains will not be rated in the study. The tool is available in the public domain.
iii. In-depth interview guide: A semi-structured in-depth interview guide will be designed to elicit the participants’ meaning and experience of recovery. The interview guide will be developed by all researchers in consultation with each other and will be content validated by 5 experts (See Table 1 for selection criteria of content validation experts). The interviews will be audio-recorded and transcribed by the researcher for subsequent analysis.
iv. Swasthya Labh Saadhan (SLS)^14^ pictorial recovery tool: The SLS is a pictorial recovery tool developed through a participatory action research method with the goal of developing a locally and contextually valid approach to understanding recovery with knowledge production that incorporates the perspectives of “experts by experience.” Developed through a partnership with the Burans organization in Dehradun, India, the themes around recovery that the tool captures through pictorial representations are: i) taking care of oneself, ii) being addiction free, iii) being spiritually engaged, iv) having fun, v) being an active family member, vi) being a friend, vii) contribute to the household, and viii) being an active community member. The tool will be administered through an interview with the participant, and permission to use the tool has been obtained from the authors.
v. Cantril’s Self Anchoring Striving Scale^24^: The Cantril self-anchoring striving scale or, commonly known as Cantril’s ladder is an instrument to measure people’s attitudes towards their life and its components in various respects. It is used to assess well-being and life satisfaction, the person’s appraisal of their life based on the best versus worst life they can imagine for themselves. Represented in the shape of a ladder that has 11 steps from 0 to 10, one’s mark on the ladder continuum represents one’s evaluation of life. The person is asked to define oneself on the continuum based on their own assumptions, perceptions, goals, and values.^24^ In the current study, only fields of present and future scores will be scored. The tool is available in the public domain and will be administered by the researcher.

#### 4. Art-based methods

To gather more insights into the subjective nature of human experience, visual art-based methods, including, but not limited to, representational forms of paintings, drawings, collages, and clay work, will be used as a method of inquiry. These interactions will be simultaneously carried out along with the participant observation and in-depth interviews and will involve a description of the art representation, its nature, why it was chosen, and how she relates to it.^25^ The method aims at bringing together the intersubjective narratives of marginalization and resilience in a space where their conversations and discussions are a part of the research process. Recognizing voices beyond languages, these sessions will allow one to express oneself through presence, participation, observation, engagement, and non-verbal interactions. 10 art-based inquiry sessions will be carried out in an open group of women with SMI, who will be requested to be a part of the group over the period of participant observations and ethnographic interactions. These women do not necessarily require to be the ones who have been interviewed.

A list of prompts will be used to guide the conversations in the group, followed by co-creating art by the researcher and participants and discussing the content of the art-why it was chosen and how they relate to it. The process of initiating the group, the dynamics of the group, and discussions in the group will be noted. In the initial two sessions of art-based inquiry, paintbrush, and watercolor/acrylics will be provided, followed by the addition of pencils, crayons, and clay in the subsequent sessions. This will be done to facilitate a free flow of art making. The prompts that will be used are presented in Table 2.

**Table 2.**
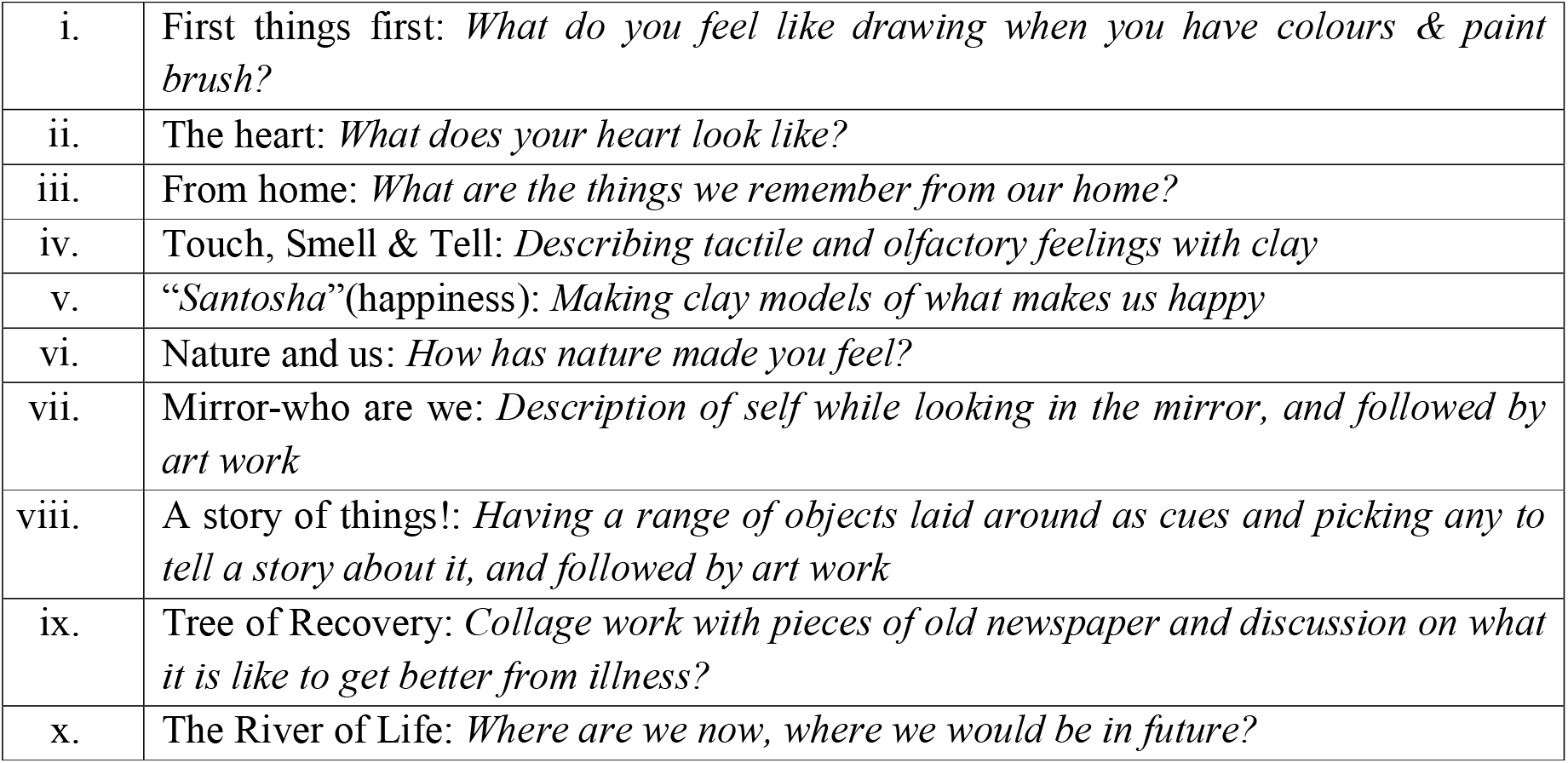
List of Prompts for art-based interaction sessions.

### Procedure

### Ethical Considerations

The study has been ethically approved by the Institutional Ethics Committee (Behavioural Sciences Division), National Institute of Mental Health and Neuro Sciences (NIMHANS), Bengaluru, India, vide reference no. NIMH/Psy/DESC Meeting-II/RP/KP/2023/22. Permission will be sought from the respective long-stay facility to collect data. All participation will be based on informed written consent, and they will be free to withdraw from the study at any stage without having to state any reason for it. Given concerns about anonymity and potential disclosure of identity, no photographs will be taken of the participants. Photographs will be taken only of the art representations they create, and the interviews will be audio recorded and transcribed; however, all information pertaining to their identity will be de-identified. No data about specific information provided by any of the participants will be shared with the treating team or administration of the long-stay facility. All identifying information will be removed while quoting verbatim from the interviews/ other data sources and for any presentation/publication. Rapport building will be emphasized to establish a relationship of trust and safety with the participants. During the observation of routine activities, if the researcher’s presence causes inconvenience or discomfort to a participant, the researcher will withdraw from the situation. In terms of researcher training, the first author has completed a 14-week online training on “Qualitative Methodology” with emphasis on Ethnography, under the supervision of Prof Michael Bamberg, Department of Psychology, Clark University, US, and former President of the APA Division 5, Society for Qualitative Inquiry in Psychology (SQIP). She has also undergone brief training with an art-therapy practitioner and mental health professional on the use and application of arts-based research methods. All the work carried out by the first author will be supervised and guided by the other three authors, each of whom has over 20 to 25 years of experience in the fields of Clinical Psychology, Psychiatry, and Medical Anthropology.

### Pilot study

A pilot study will be conducted in one long-stay facility for a duration of 10-15 days, to explore (i) the feasibility of the design, (ii) possible logistic challenges, (iii) the feasibility of contacting the centres and obtaining permission, (iv) parameters relevant or irrelevant for the observation guide; (v) applicability of the in-depth interview guide; and (vi) viability of using art-based interactions. After the pilot study, required modifications to the study design and method will be made.

### Data Analysis

Descriptive statistics using Mean, SD, and Percentages will be used to describe the sample characteristics, organization information sheet, socio-demographic and clinical data, and quantitative information on CGI, SLS and Cantril’s Ladder. This data will be analyzed using either IBM SPSS 27 Windows version or R software.

The field notes from participant observations will be analyzed using ethnographic analysis^19^ that involves (i) Reading Fieldnotes as a Data Set, (ii) Open Coding, (iii) Writing Code Memos, (iv) Selecting Themes, (v) Focused Coding, (vi) Integrative Memos, and (vii) Reflections: Creating Theory from Fieldnotes.

The in-depth interviews will be analyzed using phenomenological data analysis as outlined by Moustakas^26^ involving steps of:

i. Horizontalization-highlighting significant statements;
ii. Clusters of meaning-common meanings across the significant statements;
iii. Developing Textural descriptions-what the participants experienced, and structural descriptions-the context or setting that influenced how the participants experienced the phenomenon
iv. Composite description-reporting the common experiences of the participants, i.e., the essential, invariant structure of the data.

The art-based interactions will be analyzed through an ethno-mimetic approach as outlined by O’Neill^27^, which is particularly useful for researching marginalized or vulnerable communities through creative methods to engage with participants in ways that allow them to express their lived experiences. The output of the art-based interactions will be looked at in the context of the discourse. As an iterative and dialogic process, the group interactions will be interpreted based on the process of initiating the group, the dynamics in the group, responses to prompts, the process of co-creating art by the researcher and participants, and the content of art. The data from the multiple perspectives will be looked at through a synchronic lens, whereby the methods complement each other in understanding the accounts of recovery. It is an evolving process of research, and the possibility of finding intriguing means of interpretation remains.

## Discussion

Long-stay facilities are integral to the provision of services across a broad spectrum of mental health vulnerabilities. The study is likely to contribute towards building recovery-oriented sustainable care practices in institutional facilities. Women with SMI do not linearly progress through the illness, and experiences are context and culture-based. The findings will probably show pathways to integrate the lived experiences and meanings of recovery of women with SMI in formulating recovery-oriented care practices.

The study’s strengths include the use of diverse, in-depth lenses to understand recovery. These time-intensive and immersive observations will allow us to comprehend cultural nuances, care practices, and everyday lives in each setting. The methods are also inclusive in approach, whereby participant observations and art-based methods will help in gathering data beyond the limitations of language, speech, and symptom severity. The interviews and assessments will help in understanding individual recovery journeys and how they are situated in the context being observed.

The study holds limitations in terms of gaining access to the sites whereby research will have to follow standard procedures of each site in seeking permission for study, which can be time-consuming in itself. The time span for participant observation planned in each site could be limited to gather the vastness of data. Further, the native language of service-users may be different than that of the researcher, and communication could be challenging. The study is also limited in terms of potential observer bias, and generalizability of data.

A detailed understanding of the lived experiences and care practices for women with SMI in long-stay facilities will help us formulate recommendations that enhance their journey of recovery. Further, delving into different kinds of facilities and resource settings will build an understanding of various infrastructure and organizational practices that contribute towards care and interventions.

## Data Availability

The manuscript is a protocol for a study, and data collected subsequently shall be available upon reasonable request to the authors.

---

**Figure 1.**
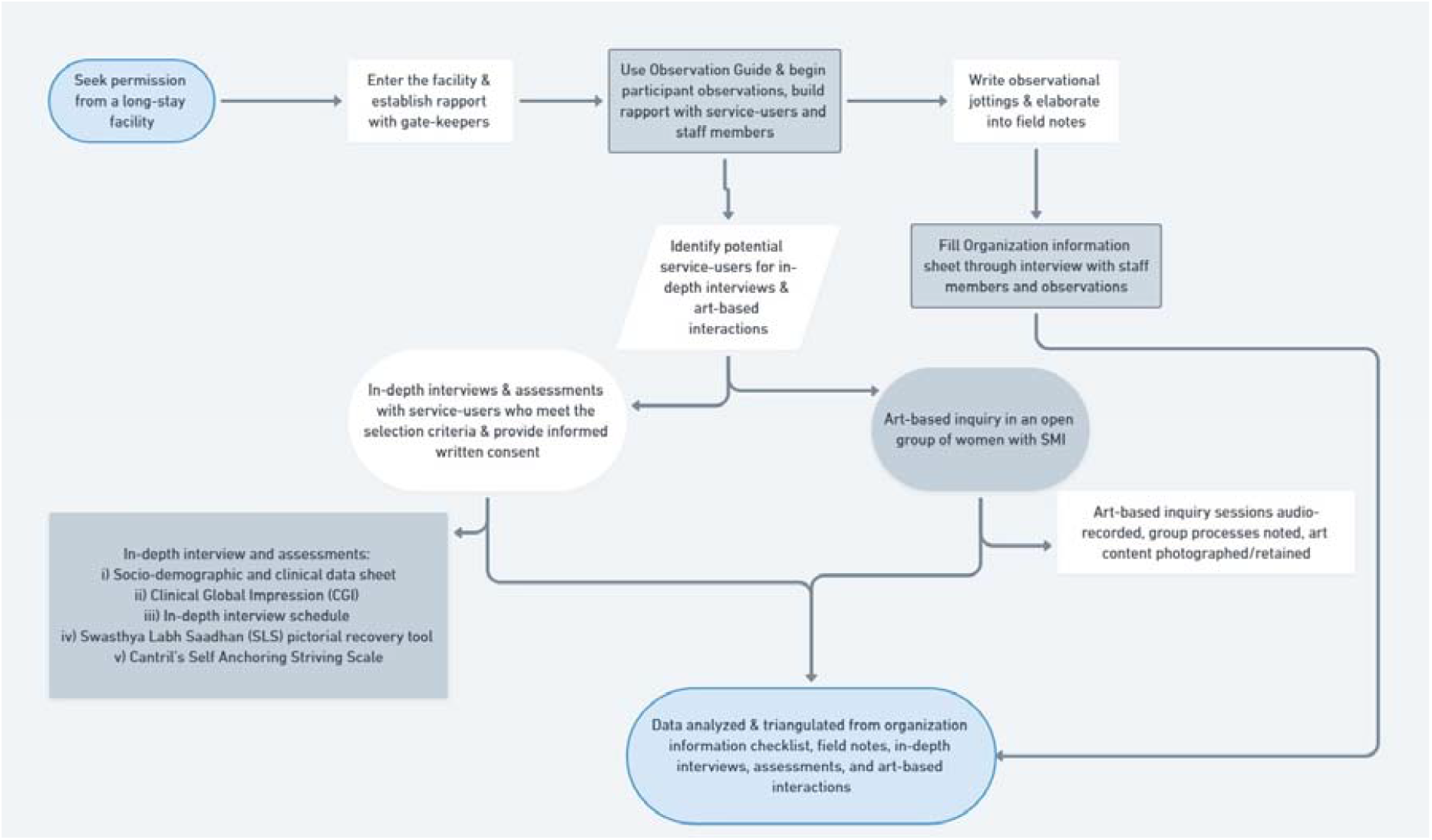
Procedure of the proposed study

## References

1. Pinto S. Daughters of parvati: women and madness in contemporary India. University of Pennsylvania Press, 2014, p.32.

2. Anthony WA. Recovery from mental illness: The guiding vision of the mental health service system in the 1990s. Psychosoc Rehabil J 1993; 16: 11–23.

3. Repper J, Perkins R. Social inclusion and recovery: A model for mental health recovery. Elsevier, 2003.

4. The World Health Organization(WHO). Mental health ATLAS 2017, https://www.who.int/publications/i/item/9789241514019 (2017, accessed January 08, 2025).

5. Murthy P, Naveen Kumar C, Chandra P, et al. Addressing concerns of women admitted to mental hospital in India: an in-depth analysis. New Delhi: National Institute of Mental Health and Neuro Sciences, Bangalore and National Commission for Women, 2016.

6. Narasimhan L, Mehta S, Ram K, et al. National strategy for inclusive and community based living for persons with mental health issues. New Delhi: The Hans Foundation, 2019.

7. National Commission for Women (NCW). Review of psychiatric homes/mental hospitals of government sector in India with special reference to the female patients in IPD. New Delhi: National Commission for Women, 2019.

8. Close H. The use of photography as a qualitative research tool. Nurse Researcher 2007; 15: 27–36.

9. Colucci E. “Focus Groups Can Be Fun”: The use of activity-oriented questions in focus group discussions. Qual Health Res. 2007; 17(10): 1422–33.

10. Department of Health, State Government of Victoria, Australia. Framework for recovery-oriented practice, https://www.health.vic.gov.au/publications/framework-for-recovery-oriented-practice (accessed January 08, 2025)

11. Daund M, Sonavane S, Shrivastava A, et al. Mental hospitals in India: Reforms for the future. Indian J Psychiatry 2018; 60(Suppl 2): S239–47.

12. NIMH. Mental Illness - National Institute of Mental Health (NIMH), https://www.nimh.nih.gov/health/statistics/mental-illness (2022, accessed January 08, 2025).

13. The World Health Organization(WHO). The ICD-10 classification of mental and behavioural disorders: Clinical descriptions and diagnostic guidelines, https://www.who.int/publications/i/item/9241544228 (1993, accessed January 08, 2025)

14. Mathias K, Pillai P, Gaitonde R, et al. Co-production of a pictorial recovery tool for people with psychosocial disability informed by a participatory action research approach—a qualitative study set in India. Health Promot Int 2020; 35(3): 486–99.

15. Anthony WA. A recovery-oriented service system: Setting some system level standards. Psychiatr Rehabil J 2000; 24: 159–68.

16. Wright PA, Kloos B. Housing environment and mental health outcomes: A levels of analysis perspective. J Environ Psychol 2007; 27(1): 79–89.

17. Kramer, MW and Adams, TE. Ethnography. In: Allen, M (ed) The SAGE encyclopedia of communication research methods. Thousand Oaks, CA: SAGE publications, Inc, 2017, pp-458–61.

18. Leavy P. Method Meets Art: Arts-based research practice. 3rd ed. Guilford publications, 2020.

19. Emerson RM, Fretz RI, Shaw LL. Writing ethnographic fieldnotes. 2nd ed. Chicago, IL: University of Chicago Press, 2011.

20. Jeffrey B, Troman G. Time for ethnography. Br Educ Res J 2004; 30(4): 535–48.

21. Eriksson P, Kovalainen A. Qualitative methods in business research London: SAGE Publications Ltd, 2008.

22. Guy W (ed). Clinical global impressions scale. In: ECDEU Assessment Manual for Psychopharmacology. Washington, DC, US Department of Health, and Welfare, 1976, pp 218–222

23. Busner J, Targum SD. The clinical global impressions scale. Psychiatry (edgmont). 2007; 4(7): 28–37.

24. Cantril H. The pattern of human concerns. New Brunswick: Rutgers University Press, 1965.

25. Guillemin M. Understanding illness: Using drawings as a research method. Qual Health Res 2004; 14(2): 272–89.

26. Moustakas C. Phenomenological research methods. Thousand Oaks California: SAGE Publications,1994.

27. O’Neill M. Asylum, migration and community. Policy Press, 2010.

